# Synthetic Data Generation for Improved covid-19 Epidemic Forecasting

**DOI:** 10.1101/2020.12.04.20243956

**Authors:** Nayana Bannur, Vishwa Shah, Alpan Raval, Jerome White

**Affiliations:** Wadhwani Institute for Artificial Intelligence; Palantir

**Keywords:** synthetic data, curve-fitting, compartmental model, SIR, COVID-19, epidemic, forecasting

## Abstract

During an epidemic, accurate long term forecasts are crucial for decision-makers to adopt appropriate policies and to prevent medical resources from being overwhelmed. This came to the forefront during the covid-19 pandemic, during which there were numerous efforts to predict the number of new infections. Various classes of models were employed for forecasting including compartmental models and curve-fitting approaches. Curve fitting models often have accurate short term forecasts. Their parameters, however, can be difficult to associate with actual disease dynamics. Compartmental models take these dynamics into account, allowing for more flexible and interpretable models that facilitate qualitative comparison of scenarios. This paper proposes a method of strengthening the forecasts from compartmental models by using short term predictions from a curve fitting approach as synthetic data. We discuss the method of fitting this *hybrid* model in a generalized manner without reliance on region specific data, making this approach easy to adapt. The model is compared to a standard approach; differences in performance are analyzed for a diverse set of covid-19 case counts.

## 1 INTRODUCTION

Infectious disease modeling plays an important role in epidemiology and public health decision making. This was apparent during the spread of covid-19, where sweeping decisions were made based on predictions from forecasting models. A challenge for modelers throughout the epidemic was providing estimates that were both sound *and* actionable: forecasts that were accurate, their parameters interpretable, and their horizons sufficient for effective policy implementation. This paper focuses on two techniques that were widely used. By themselves, the techniques excelled at subsets of those desirables. This work outlines a method for combining the techniques that succeeds at all three.

Statistical and compartmental models are popular methods for forecasting epidemics. Statistical models such as autoregressive methods and nonlinear regression attempt to model observed aspects of the disease time series. These models can be quite accurate in the short term where it is sufficient to extend a pattern of recently observed disease growth into the immediate future—a week or two, for example. Accurate longer term forecasts can be difficult to achieve because their functional forms do not generally take disease dynamics into account. This also makes it difficult to establish a correspondence between their parameters and concepts inherent to the virus or the population dynamics of the host.

Compartmental models model rates at which the virus moves through a host population. They are typically set up as systems of differential equations, capable of modeling nonlinear feedback characteristics of actual disease progression. This setup, in contrast to statistical methods, is useful for longer term epidemic understanding. Because they were built with disease progression in mind, their model parameters have disease-relevant interpretations: incubation and infectious time, or infection propensity, for example. Their structures are also flexible and can be modified to adapt to various scenarios. Coupled with parameter interpretability, this facilitates modeling various demographic breakdowns, social distancing measures, or testing policies. Because of their nonlinear dynamics, long term accuracy of compartmental models is highly dependent on accurate parameter estimation. Part of what makes that task difficult is characteristics of the data, such as small amounts in early days of an epidemic, or variability in reporting thereafter.

Together, compartmental and statistical models possess qualities that are ideal for informed policy decisions. This work, like other recent proposals [14, 16], seeks to combine these methods in a way that ultimately improves forecasting overall. We focus on two popular models in these classes: CurveFit^1^ by the Institute for Health Metrics and Evaluation (IHME) and the susceptible-infectious-removed (SIR) model well established within the epidemiological community [12]. The CurveFit framework provides a statistical model based on the Gaussian error function. It is an attractive model when disease progression is in its infancy and there are very few observed cases in an area or case precedents elsewhere. During the early days of covid-19’s spread in the United States, for example, a version of the method was heavily utilized for future case estimation. Over time, as more data became available, efforts—both at IHME and more generally—shifted from curve fitting to compartmental models [4, 10, 11]. The SIR model is the foundation on which many compartmental models stand. SIR and its extensions have been widely employed for covid-19 forecasting.

The contribution of this paper is a data-driven approach to training a compartmental model that improves its long term prediction accuracy. We find, specifically, that appending predictions from the IHME CurveFit model to ground truth observations can improve the general predictive performance of SIR models. The work examines conditions under which that improvement takes place, and outlines a technique for applying it in practice.

## 2 RELATED WORK

This work is most related to research on simulating epidemiological data, and on techniques from time series augmentation.

### 2.1 Simulated epidemic data

There have been a number of efforts to simulate covid-19 data in particular, and viral epidemic data in general. Wang et al. [18] and Doe et al. [7] develop long short-term memory (LSTM) architectures that forecast disease dynamics. In both cases, systems are trained using simulated data; Wang et al. [18] using a network simulator and Doe et al. [7] using a compartmental model. That such efforts would come from groups working with neural network models is expected, as many of those architectures require large amounts of data to properly train. The motivation behind generating simulated data is thus to generate new series of complete disease progressions as sets of independent observations. Our work is inherently different as we still rely on ground truth data during training; at least some portion of our training data is assumed to be observed rather than generated.

### 2.2 Time series augmentation

Our work is also generally related to efforts around time series augmentation. The motivation behind many of these advances is, again, to increase the amount of training data for improved system generalization. Such techniques include treating slices of a single time series as if they were unique observations, and warping portions of a time series by either compressing signal frequency or resampling [6, 9, 13]. These efforts have been applied outside of the domain of infectious disease modeling. Once applied to an epidemic curve, it is unclear what impact they would have on parameter estimation.

### 2.3 Time series forecasting

Work in time series forecasting has developed recursive-based strategies for prediction that are closely related to the methodology presented in this paper. Two established methodologies, known as *recursive* and *DirRec*, iteratively train using predicted data [1]. Unlike our methodology, the recursive strategy assumes a single model is being trained. The DirRec strategy alleviates this assumption, however, it is designed to only predict a single point into the future.

### 2.4 Combining statistical and compartmental models

The Institute for Health Metrics and Evaluation (IHME) proposed a unique combination of statistical and compartmental modeling to formulate covid-19 estimates [15, 16]. Forecasting is done in a series of steps. First, spline regression is used to make eight-day forecasts of death counts from observed data. The effective reproduction number (*R*_*t*_) is then estimated using a compartmental model (SEIR) using this extended time series. Once *R*_*t*_ is found, a regression model is used again to associate daily observations with non-epidemic covariates such as weather or mobility. That regression model is then forecast into the future to build new distributions of *R*_*t*_ estimates. Finally, those estimates are turned into long term projections using an SEIR model with initial values from the last day of the observed data.

Our methodology differs in that we are interested in the predicted values, rather than parameters estimated from those values. In this way, our hybrid model begins its estimation from the final day of short-term predicted data. More broadly, however, our motivation and subsequent approach are toward understanding the consequences of using predicted data in this paradigm. The system setup is thus simpler than the IHME one.

## 3 MODELS

This section presents the models used in our work—the IHME CurveFit model that is used as a data generator and the SIR model that is used for forecasting. The proposed *hybrid* model is an SIR model supplemented with data from CurveFit.

### 3.1 SIR (Susceptible-Infectious-Removed) model

The SIR model [12] is a well established within the epidemiological community. The model defines three compartments in which members of the host population can exist: susceptible (S), infectious (I), or removed (R). Individuals start in the S-compartment, move to infectious after coming in contact with the disease, and eventually removed once they are no longer infectious. The model is expressed as a system of differential equations that parameterize the rates at which these transitions occur:

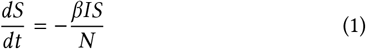

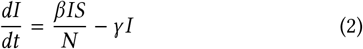

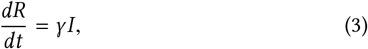

where *N* = *S* + *I* + *R*. The model has two parameters: *β* is the product of the average number of contacts per person per time and the probability of transmission during that contact; *γ* is the rate of removal from the infectious compartment. In this paper, we use the following alternate interpretation of *β* and *γ* :

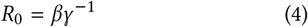

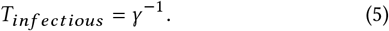

*R*_0_, the *basic reproduction number*, is the average number of secondary cases introduced due to one infected individual in a population where everyone is susceptible.

### 3.2 IHME CurveFit model

CurveFit, proposed by IHME and originally their primary method of forecasting, is a statistical model that incorporates mixed effects.

The model was designed to forecast deaths and predict the peak of the pandemic. The model fits the death rate *D*, defined as cumulative deaths per population, using the Gaussian error function:

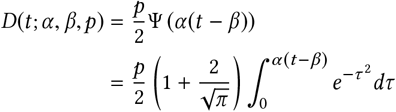

where *β* is a location-specific inflection point (time at which rate of change of *D* is maximum), *α* is a location-specific growth parameter, and *p* controls the maximum level of *D* at each location. The model was initially designed to forecast deaths. We use the model to predict total cases, but the basic premise of the model remains the same.

While the CurveFit model allows several modifications for contextualization, we choose a simple version of the model, to which we make the following assumptions: 1) only fixed effects are considered, not random effects; and 2) no covariates are used.

## 4 METHOD

This section describes the procedure by which the models outlined in the previous section (Section 3) were fit. In addition to model parameter estimation, it outlines our windowed approach to training and testing that formulated our results. The approach is motivated by reality: having a progressively updated set of observed daily infection counts and needing to make new forecasts for unseen periods.

### 4.1 Pre-Processing

Let W be a time series of observed infections. We divide W into multiple overlapping windows *W*_0:*m*_, *W*_1:*m*+1_, *W*_2:*m*+2_, …, *W*_*l* − *m*:*l*_where *m* is the length of the window and *l* is the length of the time series. Models were trained and evaluated on consecutive *W* ‘s in W. For each *W*, data for training and evaluation was created by dividing the window into two respective portions: *w*_0_, *w*_1_, …, *w*_*i*_, and *w*_*i*+1_, …, *w*_*m*−1_. The top panel of Figure 1 visualizes this process.

**Figure 1:**
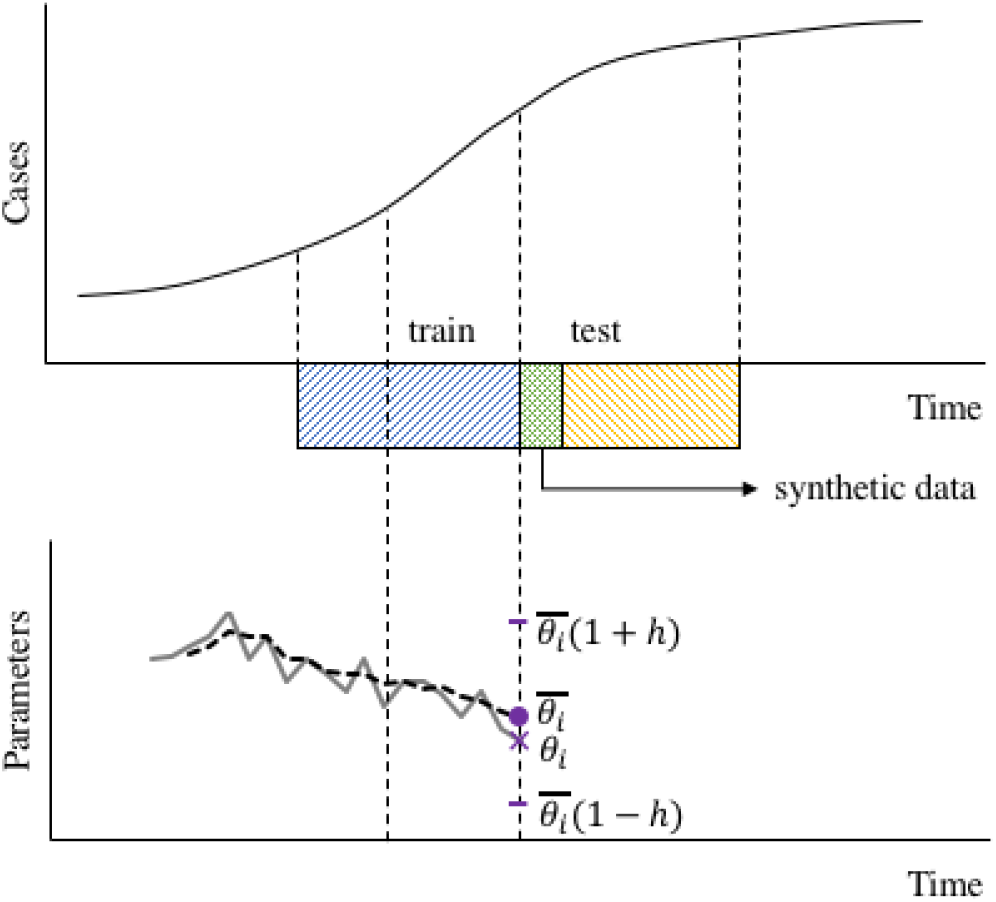
Windowed approach for training. The upper plot shows the sliding window over the time series, divided into a train set (blue) and test set (green and yellow). For the hybrid model, a portion of training data comes from CurveFit predictions (green). The lower plot shows the time series of estimated model parameters (solid). For SIR and hybrid models, a rolling mean is taken over the time series of parameters (dashes). The rolling mean at *i* is 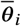. The new prior at *i* is given by 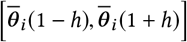. We used *h* = 0.05 in our experiments.

The act of training and evaluation for a given *W* is known as a *run*. The training portion of *W* was smoothed using a centered rolling mean with a window length of seven days. Data points lost to the rolling computation were replaced with their corresponding raw observations.

### 4.2 Fitting the SIR model

#### 4.2.1 Choice of initial conditions for SIR

The SIR model requires members of the population *N* to be assigned to one of its three compartments. Because we have actual case count estimations, this assignment can be done in an informed manner. If we assume detected cases are immediately isolated and no longer contribute to the spread of the disease, then the removed compartment (R) corresponds to the total cases reported. The removed compartment is initialized with the total cases on the day from which training starts. However, the susceptible and infected compartments are not observed. To overcome this, the initial number of infectious individuals is assumed to be some fraction of the initial total cases. That fraction, called *infectious ratio*, is treated as a latent variable and estimated in conjunction with the other model parameters. The initial number of susceptible individuals can then be found by manipulating the population assumption (Section 3.1): *S* = *N* − *R*(*I*_*r*_ + 1), where *I*_*r*_ is the infectious ratio.

#### 4.2.2 Parameter estimation

Bayesian optimization [2] was used to estimate SIR parameters, including model parameters and initial case counts. Uniform priors were specified for all parameters. The initial case count in the infectious compartment was found by estimating the infectious ratio *I*_*r*_.

The parameters of the SIR model vary depending on the data. Early in an epidemic lifecycle, there is seldom enough information to have good estimates of exact parameter values or to confidently define narrow priors that encompass an accurate value. This is further complicated by parameter differences that may arise across populations. To not bias our process to one region or point in time, we use priors with broad initial support. Table 1 details the initial parameter ranges we used irrespective of the case counts being fit.

**Table 1:** Initial ranges of model parameter distributions. All distributions are uniform, where “low” and “high” values are inclusive.

|  | SIR |  |  | IHME |  |  |
| --- | --- | --- | --- | --- | --- | --- |
| | $R_0$ | Infectious | Inf. Ratio | $\alpha$ | $\beta$ | $p$ |
| Low | 0.5 | 1.0 | 0.0 | -7.0 | 1.0 | -40.0 |
| High | 3.5 | 20.0 | 20.0 | -0.1 | 50.0 | -1.0 |

The nature of SIR models is such that several different parameter combinations can provide good fits to the underlying data. This highlights a drawback to using priors with broad support in that final parameter estimates may not be representative of realistic model parameters. While the optimization algorithm could potentially overcome this challenge, it may not do so in a reasonable time. To overcome this, we propose a historically informed fitting procedure.

The SIR model is first fit to each training window in W using the initial parameter ranges. This forms a set of best parameters *θ*_*i*_ for each window. This time series of parameters is maintained and used to inform future training. Formally, let *θ* _0_, *θ* _1_, …, *θ*_*n*_ be the time series of best-parameters found for windows *W*_0_, *W*_1_, …, *W*_*n*_. We take a three day rolling mean of the time series of parameters as shown in Figure 1. This rolling mean of parameters is denoted as 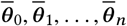 We then construct a new uniform prior 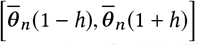 which is used to re-estimate the SIR model parameters for *W*_*n*_. Through experimentation, we fixed *h* at 0.05 for all parameters, for all training periods.

This methodology was employed because we found parameter sets obtained for consecutive runs to vary widely, more so than the incremental changes in the data would suggest. This training methodology mitigated that pattern, yielding more consistent and realistic parameter estimates across windows.

### 4.3 Fitting the IHME CurveFit model

The data fed into the CurveFit model was *log* (*X* / *N*) where *X* is the time series obtained after pre-processing (Section 4.1) and *N* is the total population of the region corresponding to the data. The CurveFit model takes bounds and initial values for its parameters as input. The uniform bounds used for the parameters are specified in Table 1. The exponential function was applied to parameters *α* and *p* to ensure that they were non-negative. The initialization of parameters was important because the optimization problem being solved is non-convex. Hence, the initial values were considered as hyperparameters and tuned by Bayesian optimization using 80 percent of the window as a training set, and 20 percent as a validation set. The model was fit within the CurveFit framework using the L-BFGS-B algorithm [3].

### 4.4 Fitting the hybrid model

The *hybrid* model is a SIR model trained using a portion of Curve-Fit’s forecast, in addition to ground truth data. Recall (Section 4.1) that training and evaluation for a window *W* take place, respectively, over points *w*_0_, *w*_1_, …, *w*_*i*_, and *w*_*i*+1_, …, *w*_*m* −1_ in *W*. For the hybrid model, the train set is extended to *w*_*i*+*n*_, where *i* +*n* < *m* and values for *i* + 1 to *i* + *n* come from estimates made by CurveFit. For this work, we fix *n* at five. This extended train set was treated as raw data—the pre-processing and SIR-specific fitting techniques previously described were applied.

## 5 EXPERIMENTAL SETUP

We performed experiments to compare the CurveFit, SIR, and the hybrid models across multiple regions and time periods. The aim of these experiments was a) to determine optimal configurations for the models used, b) to determine whether the hybrid model has a statistically significant improvement in performance over the SIR model, c) to understand how the models compare across the stages of the pandemic and across geographies, and d) to evaluate the models at various lookaheads.

### 5.1 Data setup

The datasets used consisted of publicly available daily case counts for various regions around the world. Population figures (*N*) for each region were also derived from publicly available data sources. Datasets were chosen to represent a range of geographies, administrative levels, and epidemic time scales to provide a diverse set of conditions for evaluation. Regions selected were Italy, New York City, and Delhi. The Italy data is a country level dataset, with an epidemic curve that has already peaked in the past. New York City’s case count trajectory is similar, albeit at a county level. Delhi represents state-level data but has a different case count profile. Curves for each region are visualized in Figure 2.

**Figure 2:**
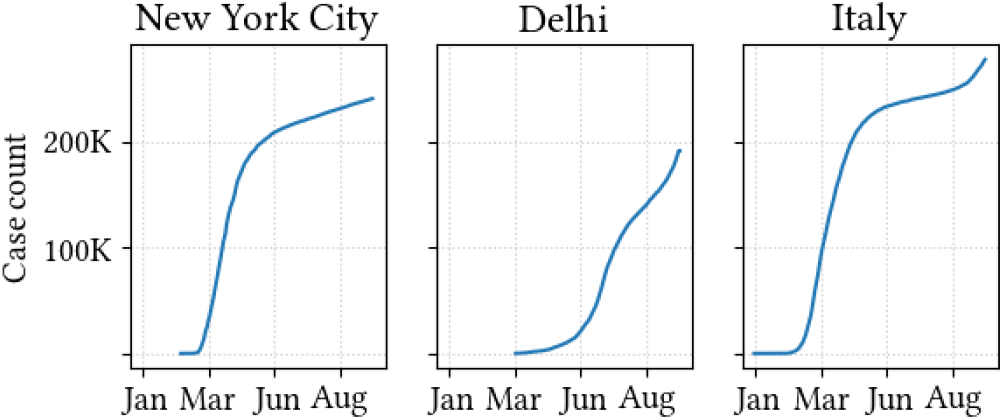
Total infected cases collected from public sources used for this analysis.

Data was downloaded from multiple sources. Data for Italy came from the Johns Hopkins Corona Virus Resource Center [8], data for New York City came from The New York Times Covid-19 Data [17], and data for Delhi came from Covid19india [5]. For each region, data was collected from the time the pandemic started in the region, or from the earliest date on which reliable data became available. For each set, the cumulative time series of total cases was used. Table 2 shows a summary of the data used.

**Table 2:** Details of case estimate data used for this study.

| Region | Start date | End date | Source | Days |
| --- | --- | --- | --- | --- |
| Italy | 19 Feb | 14 Aug | JHU | 177 |
| Delhi | 29 Mar | 18 Aug | Covid19India | 142 |
| New York City | 3 Mar | 27 Aug | NY Times | 177 |

### 5.2 Training setup

The data for each region was divided into windows as described in Section 4.1, then fit using the approaches described in Section 4.

The initial priors for each region were fixed to the defaults specified in Table 1.

An initial set of experiments was performed to determine the optimal length of the training period. In these experiments, the time series was divided into windows. For each window, the models were fit using training periods varying between six and 40 days and evaluated on a fixed 30 day test period. The models were not refitted, but multiple runs were performed for the same configuration to assess the mean and variance of the error. Based on the test MAPE, a training period of 30 days was found to be a reasonable choice across models and regions and was used in all subsequent experiments.

### 5.3 Evaluation metrics

Models were evaluated using the absolute percentage error (APE) between a given day’s estimate and its ground truth. We often aggregate APE over some dimension by taking the mean of the collection. This metric is referred to as the mean absolute percentage error (MAPE). Aggregations include all estimated days independently, estimated days within a given forecast period, and corresponding lookahead days per forecast period.

## 6 RESULTS

Recall from our experimental setup that there are several sets of training periods over a given case-count time series. For each period, the models produce estimates for several days into the future relative to that period. This section looks at the results of those estimates, aggregated in various ways. Because the first five days of forecast data were used as training for the hybrid model, evaluation starts from day six for a period of 25 days thereafter.

### 6.1 Comparison of SIR and hybrid models

The overall evaluation consists of treating each predicted day as a unique observation. Aggregation of point-wise errors in this setting is the mean over all predicted days for each model separately over every region. These means and their respective differences can be seen in the “system mean” columns of Table 3. Positive values in the difference column indicate the hybrid model having a lower MAPE than the SIR model. This was found to be the case in all regions, with differing levels of magnitude: the difference is most pronounced in Italy, with New York City having a lower, but nontrivial improvement. While there is an improvement of hybrid over SIR observed in Delhi, it is small.

**Table 3:**
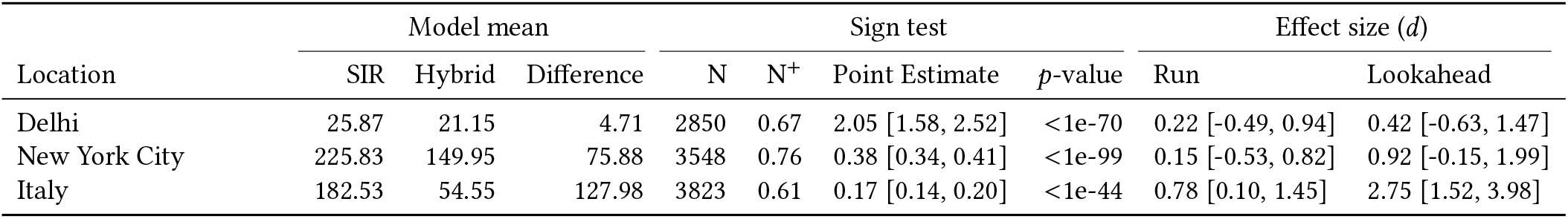
Forecast results of the SIR and hybrid models across regions. *Model mean* is MAPE observed over all forecast days; positive differences correspond to the hybrid model being better. A sign test was performed to assess whether the median of MAPE differences (*point estimate*) significantly differed from zero; N is the total number APE’s considered and N^+^ is the fraction of pair-wise differences that were greater-than zero. Effect size (*d*) is Glass’s Δ over MAPE’s aggregated per run and per lookahead day. Bracketed values are 95 percent confidence intervals around respective estimates.

To understand whether these differences were statistically significant, a sign test was performed between the difference of SIR and hybrid APEs for each region (Table 3: “sign test”). We are aware of the pitfalls around statistical hypothesis testing in this context; notably, the lack of independence across our samples (runs), and multiple hypothesis testing across regions. However, the sign test does provide a level of confidence that differences observed between models were not accidental. The median of differences in each region was found to be positive and significant at the *p* < 0.01 level. The test was set up to evaluate SIR minus hybrid, meaning a positive point estimate corresponded to a higher SIR MAPE.

### 6.2 Performance across stages of the pandemic

Instead of aggregating across all days, we now consider the MAPE grouped-by training period. This means a set of APEs aggregated per 25-day forecast window. Figure 3 presents these results, where the *x*-axis denotes the start of each training period. For each curve, the lines denote MAPE across the 25 day lookahead window that corresponds to that date’s training period. The ribbon around each curve is the 95 percent bootstrapped confidence interval of that distribution.

**Figure 3:**
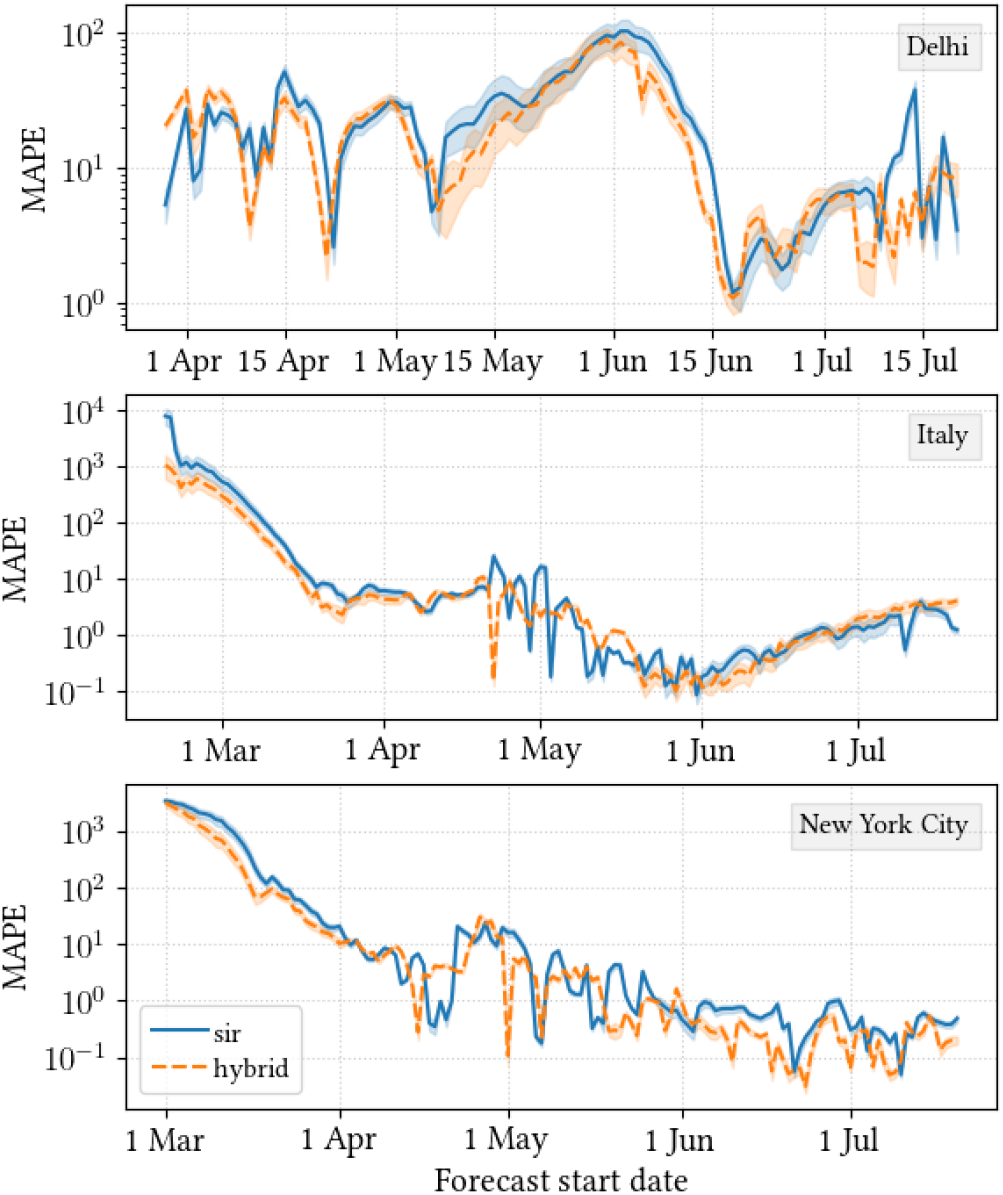
MAPE over the entire testing window, across locations.

From this perspective, we see a large benefit of the hybrid model in New York City and Italy coming from early days in the epidemic.^2^ This phenomenon is discussed in subsubsection 6.2.1. After approximately the first month, SIR and hybrid performance are similar, with the hybrid exhibiting less variance. The disease dynamics in May, for example, are difficult for SIR to consistently predict. As found from our overall assessment, the hybrid and SIR models follow one another quite closely in Delhi. In addition, the magnitude of the error overall is much less in Delhi.

Figure 4 shows differences in MAPE values between models across the epidemic time series, providing a different perspective on these results. The top figures show the observed case counts (repeated from Figure 4) while the bottom figures show the difference between hybrid MAPE and SIR MAPE. The difference is normalized by the standard deviation of SIR to be consistent with Glass’s Δ. The two sets of plots are aligned by date. The plots contain green and red highlighted regions denoting places in which the hybrid model did better or worse, respectively. A bar is present for a given day if the difference is less-than (green) or greater-than (red) one-half standard deviation across the series. Again, for New York City and Italy, we find large differences in the initial portion of the forecasts. Thereafter the training methodology favors the hybrid approach, but to a lesser extent. Delhi has an opposite pattern, and is less consistent: SIR performance is better in the beginning; the hybrid model is generally better thereafter.

**Figure 4:**
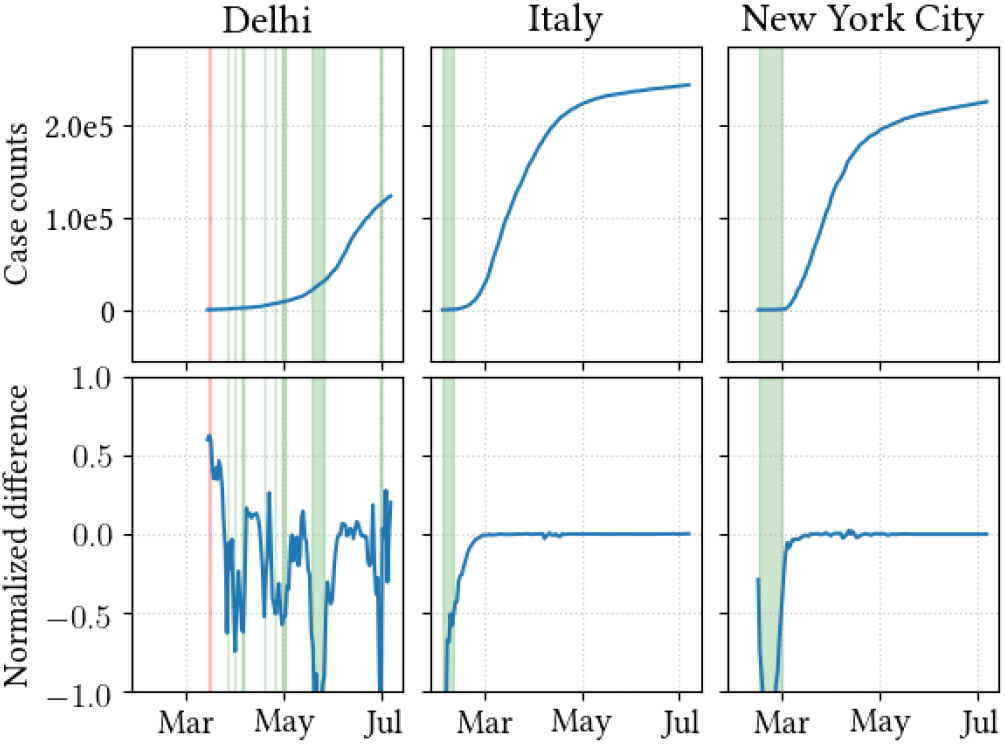
Case counts (top panels) and differences in MAPE between SIR and hybrid models (bottom panels). The normalized difference is the hybrid minus sir, divided by the standard deviation of SIR. Green bars denote regions in which the hybrid model is better-than the SIR model by more than one-half standard deviation; red bars denote the opposite.

To quantify the differences in model performance under this aggregation method, Table 3 presents effect sizes for each region (Effect size: Run). The effect size calculated is Glass’s Δ, with SIR as the control group. As other analysis has suggested, the benefit of our solution on Italian data is clear: one can expect an improvement of over one-half standard deviation on average. For New York City and Delhi, an improvement can be expected, but the magnitude is smaller.

#### 6.2.1 Performance in early phases

Evident from Figure 3 are the relatively high MAPE values early in the pandemic from both compartmental models. In New York City and Italy, the first several forecasts have MAPEs well above a few hundred. These values were maintained in our analysis because they represent a realistic situation that modelers face: having to provide forecasts when very little data is available.

Figure 5 provides an example of what is happening in this period for the Italy data set. These are plots of case counts (*y*-axis) during a subset of the time series (*x*-axis). The left-hand panels show ground truth and forecasts starting from February 20th; the right-hand panels show the same but start ten days later on March 1st. The bottom panels display the entire 25 day forecast period for actual cases, and SIR and hybrid predictions. The top panel shows the first five days of each period, and also plots CurveFit estimates for the duration.

**Figure 5:**
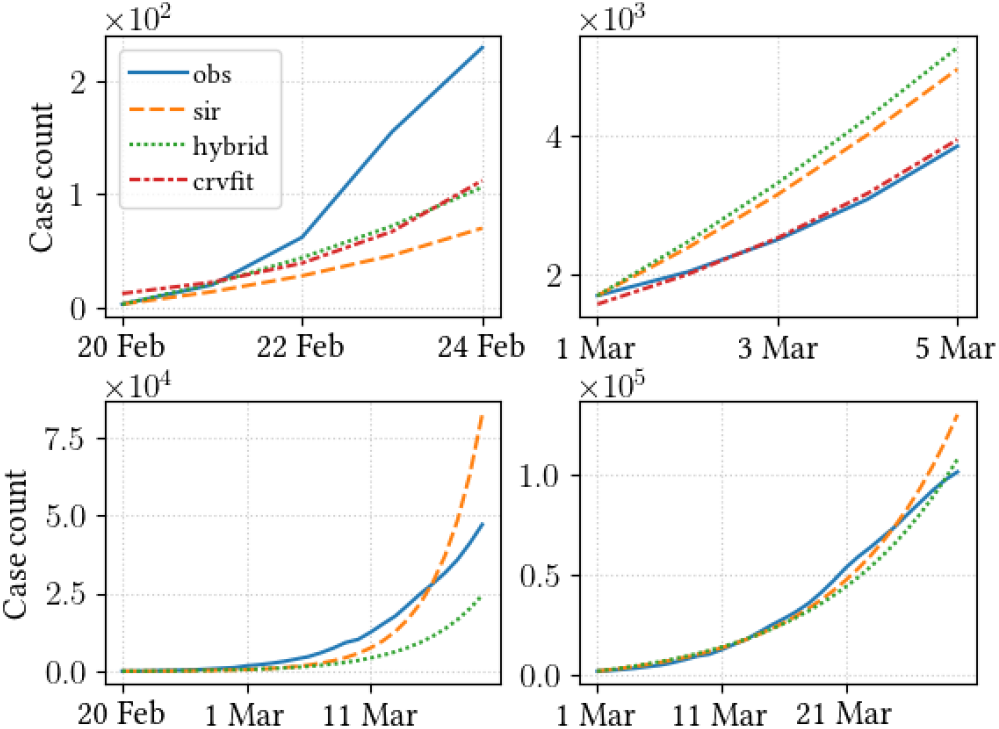
Estimated and observed (“obs”) case counts on two early days in the Italy data set. Figures on the left are for February 20; the right are for March 1. The top row displays the first five days. The CurveFit (“crvfit”) data from this period was used to train the hybrid model. The bottom row displays the 30 day estimates and observations

Rapid change in the disease progression, characteristic in the early days of these observations, results in highly dissimilar train and test sets. The parameters learned from training data are not representative of the test data. In this situation, the SIR model performs poorly as it is not equipped to handle such drastic variations. We see that in this case, CurveFit is closer to reality, using its estimates acts to bring SIR in line with that. As more consistent data forms across the test and train sets, SIR is able to get closer to the ground truth.

This situation did not occur in Delhi because the sharp exponential growth predicted by the model was not observed in that data set. This may have been in part because disease spread was contained by lockdowns, social distancing measures, and other interventions that took place earlier than the other two regions. Case growth in Delhi is thus characterized by a more gradual rise in cases in early stages than observed elsewhere. The predictions from the SIR model are far more realistic in this situation; evident from the red bars in Figure 4.

### 6.3 Performance at varying lookaheads

We now focus on average forecast-day performance for each model across various locations. Figure 6 presents a visualization. Each line represents MAPE (*y*-axis) at a given lookahead day (*x*-axis). The ribbon around each line denotes the 95 percent bootstrapped confidence interval of the distribution at that point. From this figure, the hybrid model is an improvement to the standard SIR model on just about every day, with that improvement becoming more pronounced as forecast lookahead time increases.

**Figure 6:**
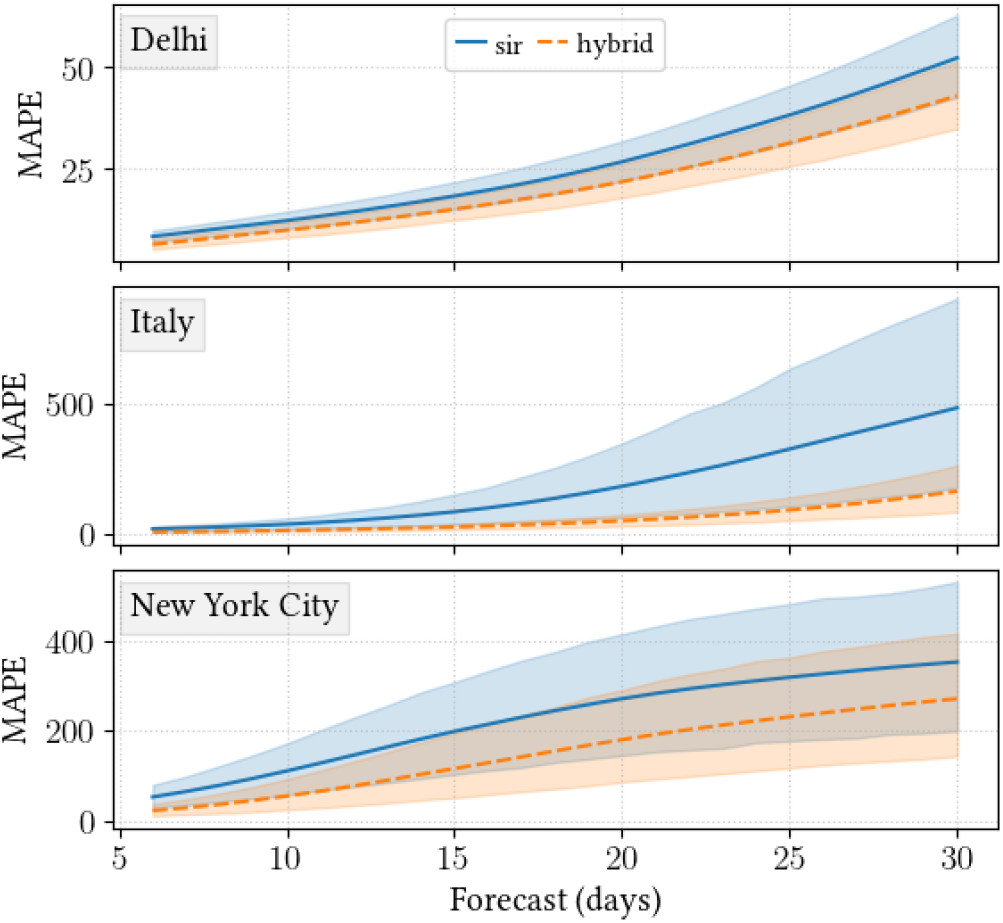
Average performance across windows for each location on every forecasted day. The windowed nature of the evaluation process produced numerous point estimates for each future-forecasted day. Days one through five were used as training for the hybrid model, and are thus not included.

Table 3 presents the effect size under this aggregation (Effect size: Lookahead). Again, we use Glass’s Δ, with SIR as the control group. The improvement seen in Italy and New York City is meaningful, with both models producing almost a standard deviation or more of improvement. The effect size in Delhi is more moderate.

## 7 THE HYBRID MODEL IN PRACTICE

There are several techniques that can be explored to improve model performance. This section looks at a rule-based approach as a first step.

### 7.1 Rule based approach

Based on analysis and exploration from Section 6, we devise a rule-based model. We have a history of evaluation data from runs that started on earlier dates, for the SIR and hybrid models. From this history, we use the results from recent runs to determine which model is likely to perform best in the future. Essentially, we choose a “best model” by comparing historical SIR and hybrid MAPEs from a subset of previous runs. While in previous sections we aggregated errors over all lookahead days to obtain the MAPE, here we aggregate errors from previous runs only over days that are in the current train window. This ensures that there is no data leakage from the forecast period. Further, since the hybrid model does not have forecasts for days one through five, we do not have evaluation data for the five most recent runs. Thus, to determine the best model for run *n*, we use evaluation data from runs *n* − 6, *n* − 7, …, *n* − *k*. Having obtained MAPEs for each of these previous runs, we further aggregate the MAPEs over all of them for each model. This results in two point estimates—historical MAPE for SIR and for the hybrid. The model with the lower historical MAPE is considered the best model and chosen for forecasting. In this situation *k* is seven, hence using seven previous runs. The value of *k* is tuned to exploit the optimal amount of past data.

### 7.2 Comparative evaluation

We test the model resulting from our rule based approach alongside the existing SIR and hybrid models, as well as three others. The first two of these three are “oracles” that are aware of model performance on the forecast day. One oracle always selects the best-performing model, while another always selects the worst. We denote these models as Ω_*b*_ and Ω_*w*_, respectively. These models implicitly establish upper and lower bounds on performance. The third model is a random selector, choosing either SIR or the hybrid with equal probability irrespective of known performance.

Table 4 presents the results of this exercise. Columns represent the various models, rows are regions in which those models were run, and values are 25-day MAPEs. None of the newly proposed models produced new forecasts. Instead, they combined existing forecasts from the SIR and hybrid models in various ways. The mean MAPEs for the SIR and hybrid models differ from Table 3 because some training periods were left out due to not being considered in the proposed model.

**Table 4:** Model performance in different regions. SIR and hybrid (HYB) models were outlined in Section 3. Other models, outlined in Section 7, generate sets of forecast by selecting forecasts from SIR and HYB. “Rand” is a random selector; “prop” is the rule based system outlined in the text. Highlighted cells denoted places in which the rule-based system had a lower MAPE than both SIR and HYB.

| Region | SIR | HYB | $\Omega_b$ | $\Omega_w$ | Rand | Prop |
| --- | --- | --- | --- | --- | --- | --- |
| Delhi | 26.64 | 20.51 | 19.77 | 27.38 | 23.53 | 21.57 |
| Italy | 32.71 | 18.11 | 17.72 | 33.10 | 24.37 | 18.06 |
| NYC | 64.32 | 29.70 | 29.09 | 64.93 | 45.88 | 29.69 |
| LA | 209.80 | 116.23 | 115.40 | 210.63 | 156.49 | 116.19 |
| Pune | 23.89 | 21.11 | 19.13 | 25.86 | 22.81 | 24.18 |

In Italy and New York City, regions in which this paper has focused, the proposed model performs closer to the best-case oracle than any other model. This is not the case in Delhi. To get a sense of its generality, the experiment was carried out in two additional regions that have thus far not been explored: Pune and Los Angeles, areas that are geographically and administratively similar to Delhi and New York City, respectively. Figure 7 plots the cumulative infected curves for these regions to provide a comparison of disease progression. Of these unseen regions, the rule based system is the best performer in Los Angeles.

**Figure 7:**
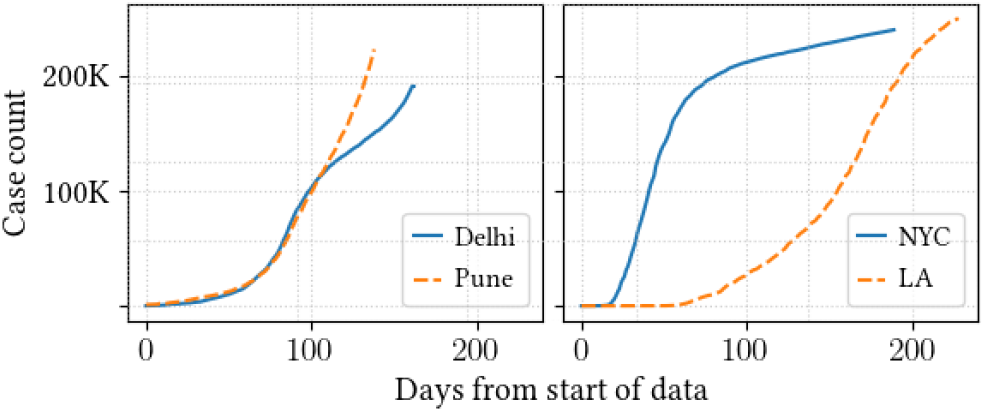
Total infected cases collected from public sources used for unseen data. Counts for seen data are also plotted for references.

While the advantage of the rule-based system is slight, differing from less than a percent on average, the results are still encouraging. Even for the regions in which the system was not better than the SIR or hybrid model, it was not far from the worst case of the two. For a practitioner, the rule-based system provides a sound methodology for using our approach. For a researcher, these results serve as a first point of comparison to alternate approaches.

## 8 CONCLUSIONS AND FUTURE WORK

This paper proposed, detailed, and evaluated a new method for training compartmental models. It was shown that by using a few days of cases forecast by a statistical model, the overall performance of the compartmental model was improved. This included not only per-day forecasts, but forecasts aggregated over the duration of the observed epidemic lifecycle. Results were shown for various regions, each exhibiting slightly different disease dynamics. Although effect sizes varied, the hybrid approach was generally better overall.

This work can be extended in a number of directions. Several training parameters were fixed, based on manual analysis of evaluation on the training sets. A natural next step is to learn these features automatically; tuning the number of days of synthetic data used, or learning a weighting of past parameter estimates to construct the prior for the present, for example. Using techniques from time series or sequence modeling could help to better estimate when and where the hybrid approach is most appropriate.

This work only considered cumulative infections. However, the hybrid model could be adapted to other use cases, such as deaths. This work could also be adapted to compartmental models more complex than SIR. Models that have additional compartments or include stochastic dynamics are more realistic, but are more difficult to train. These situations may benefit from additional future estimates.

Finally, it has been shown that covariate data—lockdowns, social distancing, variable testing rates—can have a strong impact on model performance. It could be valuable to integrate such additional data in this context.

## Data Availability

Publicly available data was used for all experiments. Data sources include (i)JHU CSSE COVID-19 Data (ii)Covid19india Data (iii)The New York Times COVID-19 Data.

https://www.covid19india.org

https://github.com/nytimes/covid-19-data

https://github.com/CSSEGISandData/COVID-19

## Footnotes

1 https://ihmeuw-msca.github.io/CurveFit

2 Note that the *y*-axis is log scale.

